# Counterfactual prediction of treatment effects on irregular clinical data using Time-Aware G-Transformers

**DOI:** 10.64898/2026.04.01.26349920

**Authors:** Gergő Hornák, Anni Heinolainen, Katalin Sólyomvári, Suvi Silén, Risto Renkonen, Miika Koskinen

## Abstract

Selecting an effective treatment relies on accurately anticipating patient’s response to alternative interventions. However, forecasting longitudinal clinical trajectories remains difficult because electronic health records contain heterogeneous, irregularly sampled data over extended time periods. These issues are especially relevant for laboratory measurements, which are central for diagnostics, assessment of therapeutic responses, and tracking disease progression in routine clinical practice. However, existing deep learning methods for counterfactual prediction usually assume regularly sampled data, an assumption incompatible with the irregular, heterogeneous data-generation processes of real-world clinical practice. Here we present the Time-Aware G-Transformer, which integrates causal G-computation with time-aware attention to predict counterfactual outcomes on irregular data. By explicitly conditioning on the timing of future observations and encoding measurement patterns, the model captures temporal dynamics that previous methods overlook. Evaluated on synthetic tumor growth data and on 90,753 cancer patient trajectories from an academic medical center, our approach demonstrates superior long-horizon (>1 day) prediction accuracy and uncertainty calibration compared to state-of-the-art baselines. These results demonstrate that embedding temporal relations directly into the attention mechanism enables robust integration of patient history data for evaluating potential treatment strategies in personalized medicine.

## 1 Introduction

In healthcare, laboratory values are often essential to clinical diagnosis and monitoring disease progression. They are also among the most frequently used parameters for assessing treatment response and toxicity over time. Estimating counterfactual treatment effects, that is, how outcomes would differ under alternative therapies, is vital for personalized medicine. However, treatment decisions depend on the patient’s current clinical state, which introduces time-varying confounding that must be properly addressed.

Statistical frameworks such as G-computation [1] and marginal structural models [2] provide principled approaches to adjusting for time-varying confounding in longitudinal data. Recent deep learning methods have extended these frameworks: G-Net [3] and G-Transformer [4] implement G-computation with neural architectures, while Causal Transformer [5] and CRN [6] use domain-adversarial training to learn treatment-invariant representations, i.e. balanced representations. These transformer-based approaches outperform traditional methods on complex, high-dimensional data [4; 5; 7], owing to their ability to capture long-range dependencies across patient histories.

However, the existing methods assume regular sampling, whereas electronic health records (EHR) and patient visits in routine practice are inherently irregular and sparse. Notably, this irregularity is informative: the timing of measurements reflects clinical decisions, such as frequent hourly monitoring in the ICU versus extended intervals of months between outpatient appointments. Thus, methods designed for regular sampling cannot capture these dynamics. Time-aware approaches such as STraTS [8], mTAND [9], and TAAT [10] handle irregular sampling through temporal embeddings, but they are designed for supervised prediction rather than counterfactual estimation, and therefore do not address the causal identification problem.

To address this gap, we propose the Time-Aware G-Transformer (TA-GT), which integrates G-computation with time-aware attention for counterfactual prediction on irregular clinical data. The key innovations of our approach are: (i) adapting the time relation mechanism based on TAAT [10] and ClinicalTAAT [11] into a causal G-computation framework, enabling counterfactual predictions that respect the temporal distance between irregularly spaced observations; (ii) reframing the causal estimand as a *joint intervention* on both treatments and measurement timings under a Measurement Timing Ignorability assumption, allowing the model to explicitly condition on future time gaps; (iii) utilizing measurement mask embeddings that encode which variables were observed at each time point, allowing the model to distinguish genuine measurements from imputed values; and (iv) integrating faithful heteroscedastic regression via Negative Log-Likelihood (NLL) optimization to capture aleatoric uncertainty, which enables the generation of well-calibrated prediction intervals.

Evaluated on synthetic tumor growth data with known counterfactuals, and on EHR data of over 90,753 long, irregular cancer patient trajectories, TA-GT demonstrates superior long-horizon prediction accuracy compared to baseline models.

## 2 Problem Formulation

We predict time-varying continuous outcomes over an arbitrary prediction horizon given irregular, sparse longitudinal EHR history. For each patient *i* ∈ {1, …, *N*}, we observe a sequence of clinical events at times 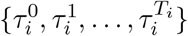, where *T*_*i*_ denotes the total number of events in the sequence of a patient. At each time step *t*, the patient state consists of:

- a vector of continuous covariates 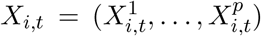, where *p* is the number of covariates, from which 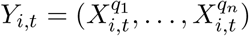 are the target outcomes to be predicted where {*q*_1_, …, *q*_*n*_} ⊂ {1, …, *p*},
- a measurement mask 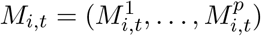 indicating which covariates are observed 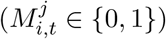,
- a vector of binary treatment assignments 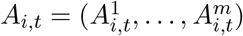, where *m* is the number of treatments,
- time-fixed contextual features (e.g., demographics) 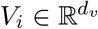.

The elapsed time between consecutive observations is denoted as 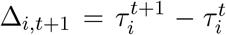. The full observed history up to time *t* is defined as

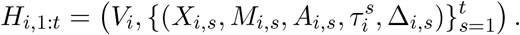

Throughout this paper, we use uppercase letters (e.g., *A*_*t*_, Δ_*t*_) to denote random variables and lowercase letters (e.g., *a*_*t*_, *δ*_*t*_) to denote their specific realizations. In standard EHR settings, the time between visits (Δ) is a clinical decision that may be influenced by prior treatments and the patient state, potentially acting as a post-treatment collider. To ensure causal identifiability, we define the counterfactual estimand under a **joint intervention** that sets both the treatment strategy and the future monitoring schedule. Given the history, the goal is to predict the distribution of the next observation under a specific treatment *a*_*t*_ and time gap *δ*_*t*+1_:

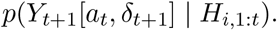

This formulation is extended to a multi-step counterfactual trajectory prediction under a specific treatment strategy *ā*_*t*:*t*+*τ*−1_ and a scheduled sequence of future time gaps 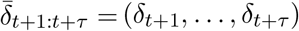, where the sequence of random variables for future treatments is denoted as *Ā*_*t*:*t*+*τ* −1_ = (*A*_*t*_, *A*_*t*+1_, …, *A*_*t*+*τ* −1_):

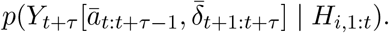

By treating the time gaps as part of the hypothetical intervention, we explicitly condition on future measurement timings 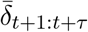 without introducing collider bias. This decouples the question of what the outcome will be from when it will be observed, enabling the clinically relevant question: *What would the patient’s state be if they received this treatment regimen and were followed up at these specific time points?* This yields future observation times 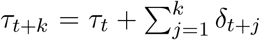, enabling predictions at clinically meaningful timepoints.

The model estimates the conditional distribution of the outcomes. For benchmarking, we evaluate point predictions: 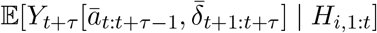. For notational convenience, later when discussing a single patient trajectory, we omit the patient index *i*.

## 3 Methods

### 3.1 G-Computation

G-computation [1] provides a direct approach to estimate counterfactual outcomes by modeling the data-generating process. To identify the joint intervention estimand defined in section 2, we rely on four causal assumptions. *Consistency* requires that observed outcomes equal counterfactual outcomes under the received treatment and monitoring schedule. *Positivity* requires that each treatment and time gap has non-zero probability given any history. *Sequential ignorability* requires that treatment assignment is independent of future potential outcomes given observed history. Moreover, we extend G-computation by assuming *Measurement Timing Ignorability*: the timing of the next measurement Δ_*k*+1_ is conditionally independent of unobserved potential outcomes, given the fully observed past patient history.

Under these assumptions, the counterfactual distribution can be identified via the G-formula. Because the target outcomes *X*_*t*+*τ*_ are a subset of the full state vector *X*_*t*+*τ*_, we first model the joint counterfactual distribution of *X*_*t*+*τ*_ and then extract the relevant components for *Y*_*t*+*τ*_. Let 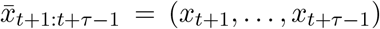 denote an intermediate state trajectory and 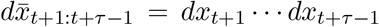. Conditioning on a specified sequence of future time gaps 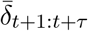 gives:

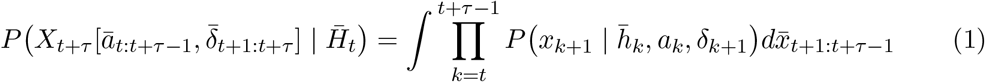

where 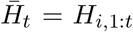 is the initially observed history up to time *t*, and 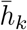 represents the autoregressively constructed history up to step *k*, formed by recursively appending the simulated state *x*_*k*_, treatment *a*_*k*_, and time gap *δ*_*k*_ to 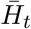. The marginal counterfactual distribution of *Y*_*t*+*τ*_ is then obtained by extracting the relevant dimensions from *X*_*t*+*τ*_ .

This formula marginalizes over all possible intermediate covariate trajectories 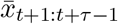 under the joint treatment strategy *ā* and the scheduled time gaps 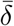, weighted by their probabilities given the history. In practice, calculating this integral directly is not feasible with high-dimensional continuous covariates. Instead, deep learning models such as G-Transformer [4] and our proposed TA-GT approximate it via Monte Carlo simulations. Counterfactual trajectories are generated by autoregressively sampling from the learned one-step-ahead conditional distributions 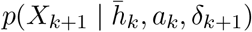, detailed below. This simulation process is performed repeatedly for each instant *t*, producing an empirical distribution of the counterfactual outcomes.

### 3.2 Model Architecture

#### 3.2.1 Input Representation

Each patient’s longitudinal data is mapped into embeddings representing continuous covariates, measurement masks, treatments, and event times.

At each timestamp *τ*_*t*_, the model receives (i) mean and standard deviation (z-score) normalized continuous covariates *X*_*t*_ ∈ ℝ^*p*^, where unobserved values are imputed with the population mean (zero); (ii) the corresponding binary measurement mask *M*_*t*_ ∈ {0, 1}^*p*^ indicating which values were observed; (iii) binary treatment assignments *A*_*t*_ ∈ {0, 1}^*m*^; static demographic features *V*; and (v) timestamp decompositions *h*(*τ*_*t*_) (e.g., year, month, day, hour).

Input features are projected into a common embedding space using separate subnetworks:

- **Continuous Covariates:** Each of the *p* covariates is passed through its own MLP, and the results are summed: 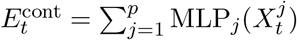.
- **Measurement Mask:** The mask vector is projected with a linear layer: 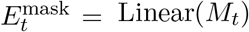.
- **Treatments:** Each of the *m* treatments has a corresponding embedding vector, which are summed: 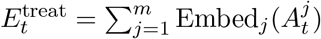.
- **Demographics:** Static features are projected via an MLP: 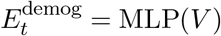.
- **Time:** Hierarchical time features are embedded and summed: 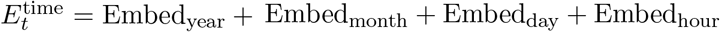.

The final input embedding for the transformer at time *t*, 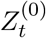, is created by summing the relevant component embeddings:

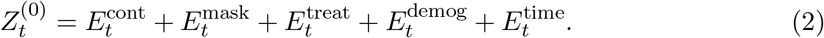

Unlike TAAT [10], which uses time embeddings only for computing the attention bias, TA-GT additionally incorporates the time embeddings into the input representation via 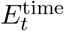. This allows the model to encode temporal context at both the representation and attention levels, yielding richer hidden states.

#### 3.2.2 Time-Aware Attention Mechanism

To model irregular sampling, we adopt the Time Relation Estimation (TRE) mechanism from TAAT [10]. TRE encodes pairwise temporal relationships between all observations into a bias matrix that modifies the self-attention scores, allowing the model to up-weight or discount information based on temporal proximity. From the time embeddings 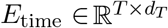, the time relation matrix *R*^∗^ ∈ ℝ^*T* ×*T*^ is computed as:

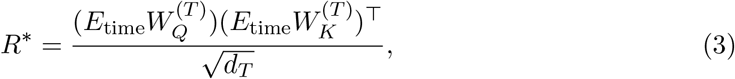

where 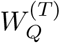 and 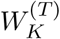 are learnable projection matrices. This matrix encodes pairwise temporal dependencies between observations and serves as an additive bias in the scaled dot-product attention:

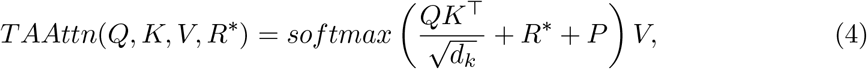

where *P* denotes a causal mask preventing access to future tokens.

The time relation matrix is incorporated when computing attention heads, which is noted for the *n*th head as:

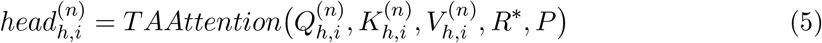

The multi-head attention is calculated as:

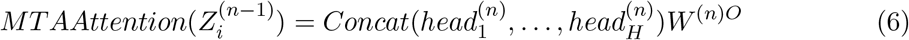

Then the output of the transformer block is

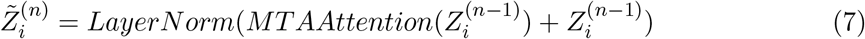

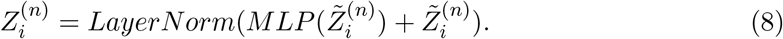

##### Conditioning on the next measurement time

To predict *X*_*t*+1_ at the next times-tamp *τ*_*t*+1_, we additionally embed *h*(Δ_*t*+1_) and project it to the hidden dimension:

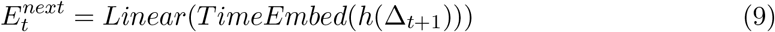

Then we add it to the last hidden state, which will encode the information about the time of the next sampling point:

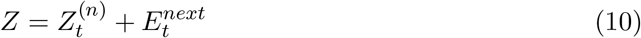

##### Prediction heads

After obtaining the final hidden state *Z*, we can use it to predict the next measurement *X*_*t*+1_ through different prediction heads for each covariate. Each *Head*_*k*_ denotes a linear layer.

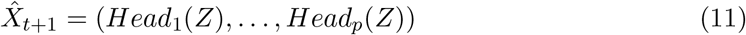

#### 3.2.3 Training AND Counterfactual Simulation

The main optimization goal of TA-GT is to predict time-varying outcomes *Y*. During training, the model is optimized on all covariates *X* using teacher forcing: at each step *t*, the model receives the ground truth past sequence ℋ_*i*,1:*t*_, denoting the structured history of covariates, masks, treatments, and timestamps, to predict the next observation *X*_*t*+1_. A masked loss is applied only to measured variables *j* where 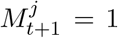. The loss function consists of MSE for point prediction and negative Log-Likelihood (NLL) for distributional prediction (Appendix B).

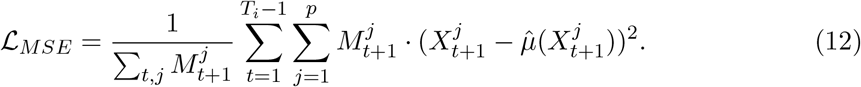

For counterfactual simulation under a given treatment strategy and specified future time intervals 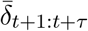, the trained model autoregressively generates trajectories 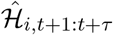 using Monte Carlo simulations. At each future step *k* from *t* + 1 to *t* + *τ*, the treatment from the strategy and the specified time intervals are applied, and the next state 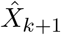 is predicted based on the history *Ĥ*_*i*,1:*k*_. Noise is then added to the predicted covariates to approximate the distributions in the G-computation integral. With MSE training, noise is sampled from empirical one-step-ahead residuals 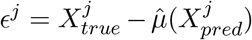 which are computed on a held-out validation set.

## 4 Experiments

We assessed the models using two datasets: synthetic tumor growth data, which offers ground truth counterfactuals for evaluating causal performance, and a real-world EHR cohort to test the factual prediction of laboratory values from irregular patient histories. Each experiment is repeated with varying data sparsity to assess robustness in conditions akin to real-world scenarios.

### 4.1 Model evaluations with Synthetic Data

To establish ground truth for counterfactual trajectories, we employed pharmacokinetic-pharmacodynamic (PK-PD) modeling [12], previously used in [4; 5], simulating tumor volume (in mm^3^) as the continuous outcome. The tumor volume is influenced differently by binary chemotherapy and radiotherapy treatments. At each time step, treatment could be administered or withheld, and the outcome was observed.

To test the models’ capability in handling irregularity, we introduced a controllable sparsity mechanism. The probability of observing a measurement on a given day is higher near treatments, mimicking clinical practice where patients are monitored more closely during interventions. On regular days, measurements were recorded with base probability *p*. On the treatment day, measurements were always recorded, and for the next three days, after the treatment, the probability of observation increased to 3*p*. We evaluated four sparsity levels: {1.0, 0.3, 0.1, 0.02}, corresponding to expected observation intervals of approximately 1, 3, 10, and 50 days, respectively. Patient sequences contained up to 60 time steps, unless terminated earlier due to tumor exceeding a fatal threshold or recovery. As sparsity increases, patient trajectories span longer calendar periods and contain more treatment events. Models were trained on 10,000 patient trajectories with 1,000 for validation and 1,000 for testing. Predictions were evaluated starting from time step 50.

We evaluated RMSE in two scenarios. In the **Randomized Counterfactual** setting, treatments were assigned at random (probability 0.1) during the prediction horizon (1–10 steps), regardless of patient history, to assess unbiased outcome prediction. For the **Fixed Counterfactual** setting, we compared four strategies over a 1–4-step horizon: no treatment, chemotherapy only, radiotherapy only, and combined therapy.

### 4.2 Real-World Data

We evaluated all models on an EHR cohort from Helsinki University Hospital (HUS), comprising 90,753 cancer patients with follow-up periods of up to five years from the time of diagnosis. Unlike the one-dimensional synthetic dataset, the EHR data included 49 common laboratory measurement types (e.g., creatinine, hemoglobin, electrolytes), binary indicators for drug administrations defined by ATC codes, and static demographic information such as age at diagnosis, gender, and cancer type. To assess the model performance, we evaluated the impact of propionic acid derivatives (M01AE), which are recognized for their influence on renal function, on laboratory test results. Continuous values were normalized to zero mean and unit standard deviation. At each time point where some medical event was recorded, missing lab values were imputed with zero, with their mask embeddings also set to zero. Time was measured in hours since diagnosis, with TA-GT decomposing it into year, month, day, and hour.

Since counterfactual ground truth is unavailable in real data, we evaluated factual prediction accuracy: predicting the future laboratory values given the longitudinal patient history by taking the mean of 50 Monte Carlo simulations at each timestep for all laboratory values. We used patient histories up to 300 events, covering 97.8% of patients whose sequences contained at most 300 events which is shown in Figure D1. The data was split 80%, 10%, 10% for training, validation, and testing.

The study was approved by HUS Helsinki University Hospital (permit HUS/223/2023); no further ethical permission or informed consent was needed for retrospective registry data under national and EU law. Data storage and analysis took place on a secure analytics platform (HUS Acamedic), maintaining patient confidentiality and complying with the Finnish Medical Research Act (552/2019) for secondary use of medical records.

### 4.3 Model Comparisons

We compared TA-GT against two state-of-the-art transformer-based models: G-Transformer (GT) [4], which implements G-computation with standard transformers, and Causal Transformer (CT) [5], which uses balanced representations. Since these baselines were designed for regular sampling and thus did not incorporate time-embeddings, we adapted them using Time2Vec [13] embeddings, next-time-delta conditioning, and measurement mask embeddings (Appendix A). Hyperparameter tuning was performed separately for each model using random search [14] on the validation set. All models were implemented in PyTorch and trained on NVIDIA T4 GPUs.

## 5 Results

We present results comparing TA-GT against the baseline models on both synthetic and real-world data.

### 5.1 Counterfactual prediction using synthetic data

We first assessed counterfactual prediction using the synthetic dataset, where the outcomes of alternative treatments were known. Figure 2a shows TA-GT’s factual predictions with 90% confidence intervals. Figure 2b shows counterfactual predictions.

**Figure 1:**
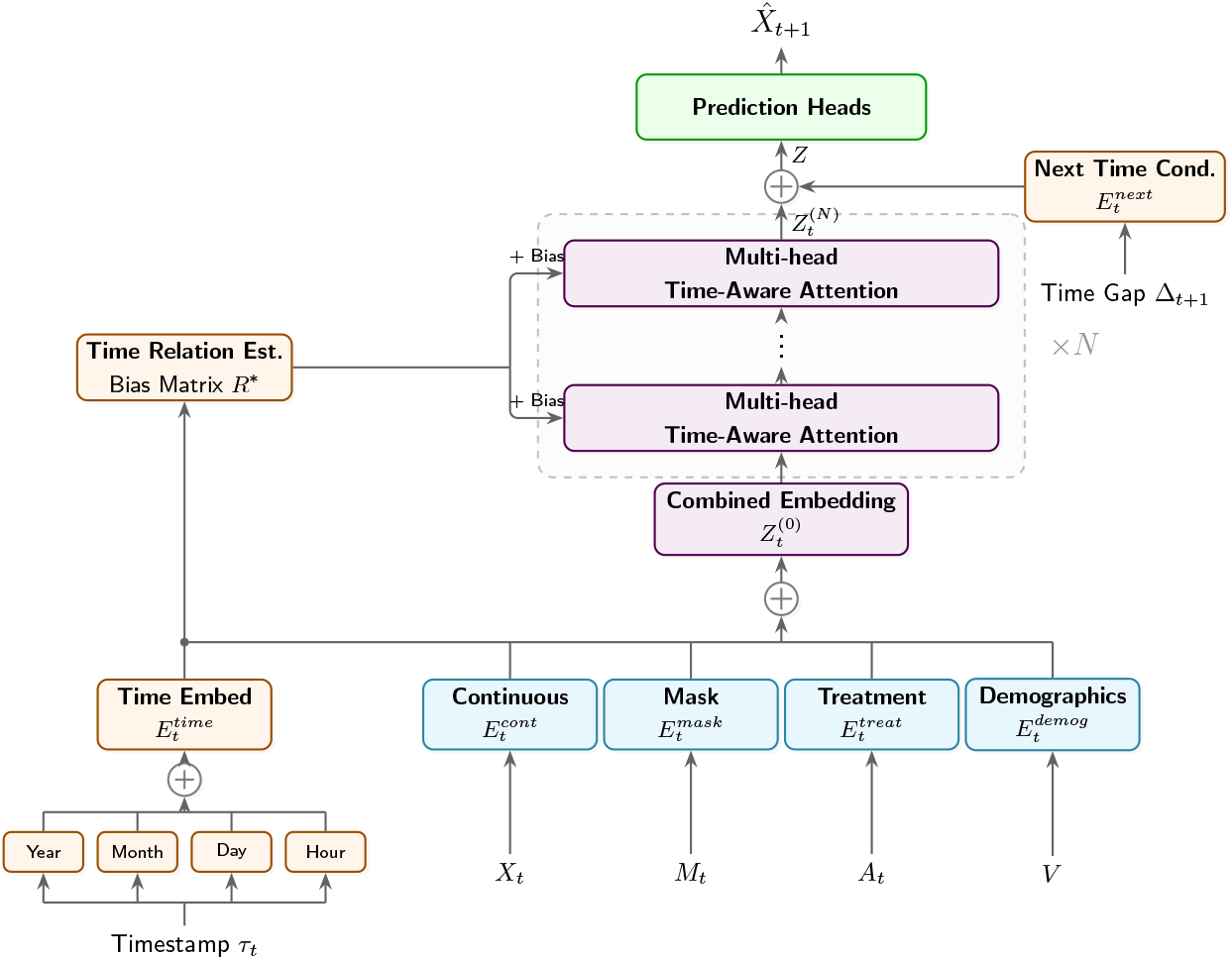
Overview of the proposed architecture. Combining different input features into a unified embedding which is processed by N transformer layers. The attention mechanism is modified with the time relation estimation bias matrix. Predictions are conditioned on future time gaps.

**Figure 2:**
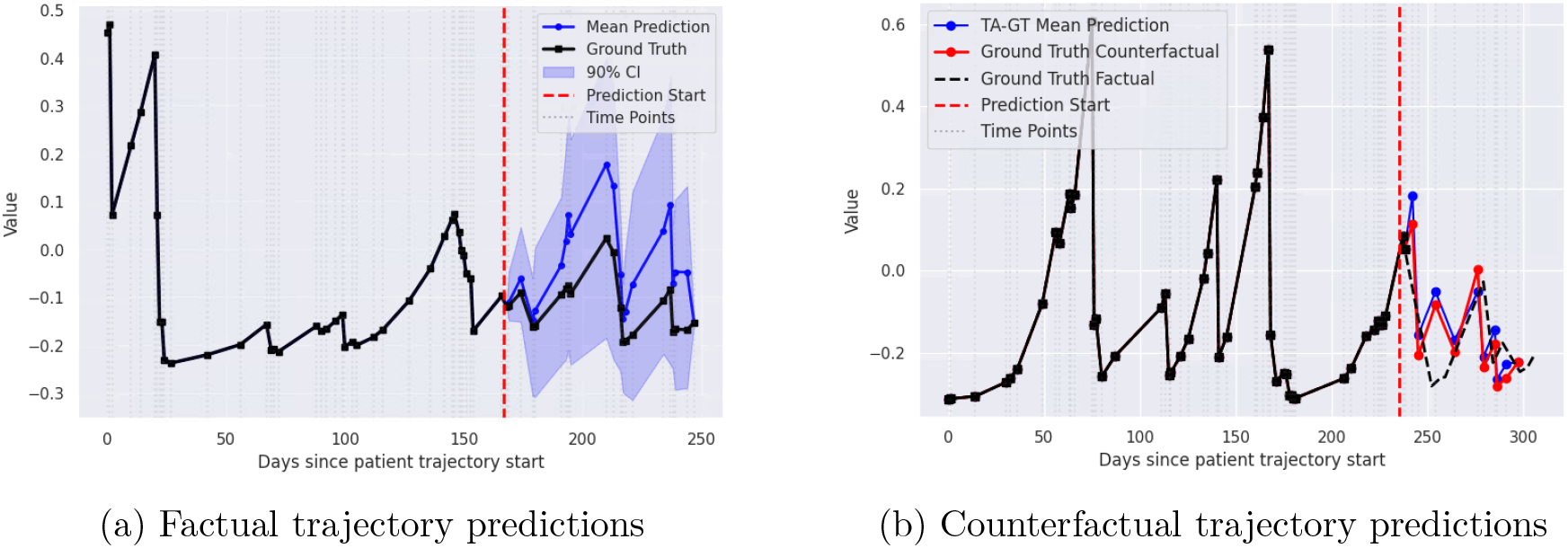
TA-GT predictions on (normalized) synthetic data (sparsity 0.1). (a) Factual predictions with 90% CIs for a 20 time step horizon based on 40 observed events. (b) Counterfactual predictions. The vertical dashed line marks the start of the prediction horizon, after which the model generates predictions autoregressively. Colors show predicted outcomes under different treatment strategies.

Table 1 summarizes prediction performance for the randomized counterfactual setting across ten consecutive time steps and different degrees of data sparsity. As the sparsity controlling variable decreased, performance degraded for all models due to increased uncertainty. TA-GT consistently outperformed both baselines, especially at shorter prediction horizons.

**Table 1:** Prediction performance on synthetic data. Tumor volume RMSE (mm^3^) under randomized treatment assignment during prediction horizon. Lower is better.

| Model | Sparsity | Step 1 | Step 2 | Step 3 | Step 4 | Step 5 | Step 6 | Step 7 | Step 8 | Step 9 | Step 10 |
| --- | --- | --- | --- | --- | --- | --- | --- | --- | --- | --- | --- |
| **CT** | 1 | 1.83 | 2.84 | 3.81 | 4.82 | 5.68 | 6.34 | 6.91 | 7.51 | 8.28 | 9.03 |
| **GT** | 1 | 0.87 | 1.52 | 2.13 | 2.64 | 3.20 | 3.81 | 4.36 | 4.97 | 5.54 | 6.10 |
| **TA-GT** | 1 | **0.61** | **0.96** | **1.22** | **1.47** | **1.78** | **2.08** | **2.39** | **2.68** | **3.04** | **3.50** |
| **CT** | 0.3 | 2.33 | 4.05 | 5.42 | 7.25 | 8.12 | 9.73 | 11.97 | 12.78 | 15.66 | 19.38 |
| **GT** | 0.3 | **1.27** | 2.03 | **3.30** | 4.27 | 5.68 | 6.90 | **7.71** | **10.00** | 14.26 | **14.12** |
| **TA-GT** | 0.3 | 1.32 | **1.98** | 3.30 | **4.06** | **5.53** | **6.70** | 7.86 | 10.31 | **10.51** | 14.51 |
| **CT** | 0.1 | 3.45 | 5.81 | 9.15 | 13.73 | 15.34 | 18.32 | 21.69 | 25.64 | 31.91 | 34.79 |
| **GT** | 0.1 | 3.00 | 5.11 | 7.33 | 10.82 | 15.37 | 17.57 | 20.10 | 23.61 | 27.61 | 34.03 |
| **TA-GT** | 0.1 | **2.79** | **4.77** | **6.86** | **9.82** | **14.29** | **16.24** | **18.91** | **22.02** | **25.80** | **31.95** |
| **CT** | 0.02 | 10.09 | 20.38 | 40.61 | 51.17 | 61.22 | **49.96** | 59.53 | 76.06 | **78.30** | **80.45** |
| **GT** | 0.02 | 4.50 | 13.20 | 23.09 | **43.07** | 54.88 | 64.26 | 54.95 | 64.70 | 81.41 | 83.39 |
| **TA-GT** | 0.02 | **4.18** | **12.72** | **22.84** | 43.09 | **54.82** | 64.28 | **54.29** | **63.72** | 80.47 | 82.27 |

Table 2 shows performance under four different treatment strategies applied over consecutive time steps. TA-GT performs best on dense data. At extreme sparsity (0.02), CT occasionally outperforms TA-GT.

**Table 2:** Prediction of tumor volume RMSE (mm^3^) under fixed treatment strategies (no treatment, chemotherapy, radiotherapy, or both) applied for 1-4 consecutive steps. Lower is better.

| Treatment | Step | Sparsity: 1 |  |  | Sparsity: 0.3 |  |  | Sparsity: 0.1 |  |  | Sparsity: 0.02 |  |  |
| --- | --- | --- | --- | --- | --- | --- | --- | --- | --- | --- | --- | --- | --- |
|  |  | CT | GT | TA-GT | CT | GT | TA-GT | CT | GT | TA-GT | CT | GT | TA-GT |
| **No treatment** | 1 | 1.63 | 0.83 | **0.38** | 1.81 | 0.89 | **0.77** | **1.48** | 1.90 | 1.88 | **1.79** | 4.33 | 3.80 |
|  | 2 | 2.96 | 1.51 | **0.67** | 3.49 | 1.78 | **1.55** | **2.86** | 3.58 | 3.99 | **3.51** | 8.90 | 8.30 |
|  | 3 | 4.26 | 2.29 | **0.97** | 5.02 | 2.91 | **2.45** | **4.23** | 5.16 | 6.40 | **5.18** | 14.70 | 13.72 |
|  | 4 | 5.33 | 2.89 | **1.26** | 6.48 | 4.11 | **3.50** | **5.57** | 6.62 | 9.09 | **6.82** | 21.30 | 19.51 |
| **Chemo** | 1 | 2.28 | 0.90 | **0.45** | 1.43 | 0.68 | **0.42** | **1.02** | 1.61 | 1.14 | **1.15** | 2.86 | 2.90 |
|  | 2 | 4.18 | 1.35 | **0.76** | 2.73 | 1.06 | **0.96** | **1.55** | 2.05 | 1.83 | **1.74** | 5.34 | 5.84 |
|  | 3 | 5.73 | 1.70 | **1.24** | 3.89 | **1.56** | 1.56 | **2.15** | 2.66 | 2.78 | **2.24** | 7.61 | 8.77 |
|  | 4 | 6.87 | 2.00 | **1.73** | 4.79 | **1.96** | 2.02 | **2.75** | 2.93 | 3.39 | **2.70** | 9.36 | 11.19 |
| **Radio** | 1 | 4.00 | **2.23** | 2.31 | 4.12 | 1.76 | **1.52** | 3.31 | 1.90 | **1.72** | 2.63 | **2.56** | 3.16 |
|  | 2 | 4.50 | 2.88 | **2.61** | 5.32 | 2.32 | **1.90** | 4.08 | 2.68 | **2.17** | **3.28** | 3.38 | 5.23 |
|  | 3 | 4.44 | 3.25 | **2.81** | 5.90 | 2.60 | **2.09** | 4.47 | 2.93 | **2.84** | **3.52** | 3.86 | 7.20 |
|  | 4 | 4.39 | 3.55 | **3.23** | 6.09 | 2.94 | **2.27** | 4.63 | 3.09 | **3.05** | **3.59** | 4.20 | 8.53 |
| **Both** | 1 | 5.81 | 2.39 | **2.34** | 3.97 | 2.45 | **1.67** | 5.06 | 2.24 | **1.87** | 3.93 | **2.66** | 3.27 |
|  | 2 | 6.57 | 2.11 | **2.10** | 4.22 | 2.63 | **1.61** | 4.10 | 2.28 | **1.95** | **2.85** | 2.90 | 4.38 |
|  | 3 | 6.56 | **1.55** | 1.67 | 3.64 | 2.36 | **1.50** | 2.81 | 2.19 | **1.96** | **2.86** | 2.87 | 5.65 |
|  | 4 | 6.73 | **1.85** | 2.25 | 3.30 | 2.22 | **1.42** | 2.69 | **1.95** | 2.03 | **2.03** | 2.97 | 5.99 |

### 5.2 Real-World Data Performance

#### 5.2.1 Predictive Accuracy

We evaluated factual prediction accuracy over a ten-step prediction horizon using normalized RMSE. Figure 3 presents results for plasma creatinine. TA-GT outperformed both baselines at all history lengths and horizons, with the performance gap increasing at longer horizons. With 100-event histories, TA-GT’s 10-step RMSE (0.37) was 41% lower than GT’s (0.63) and 58% lower than CT’s (0.89), showing roughly a 17 *µ*mol/l improvement. TA-GT’s error remained stable, whereas baseline errors increased.

**Figure 3:**
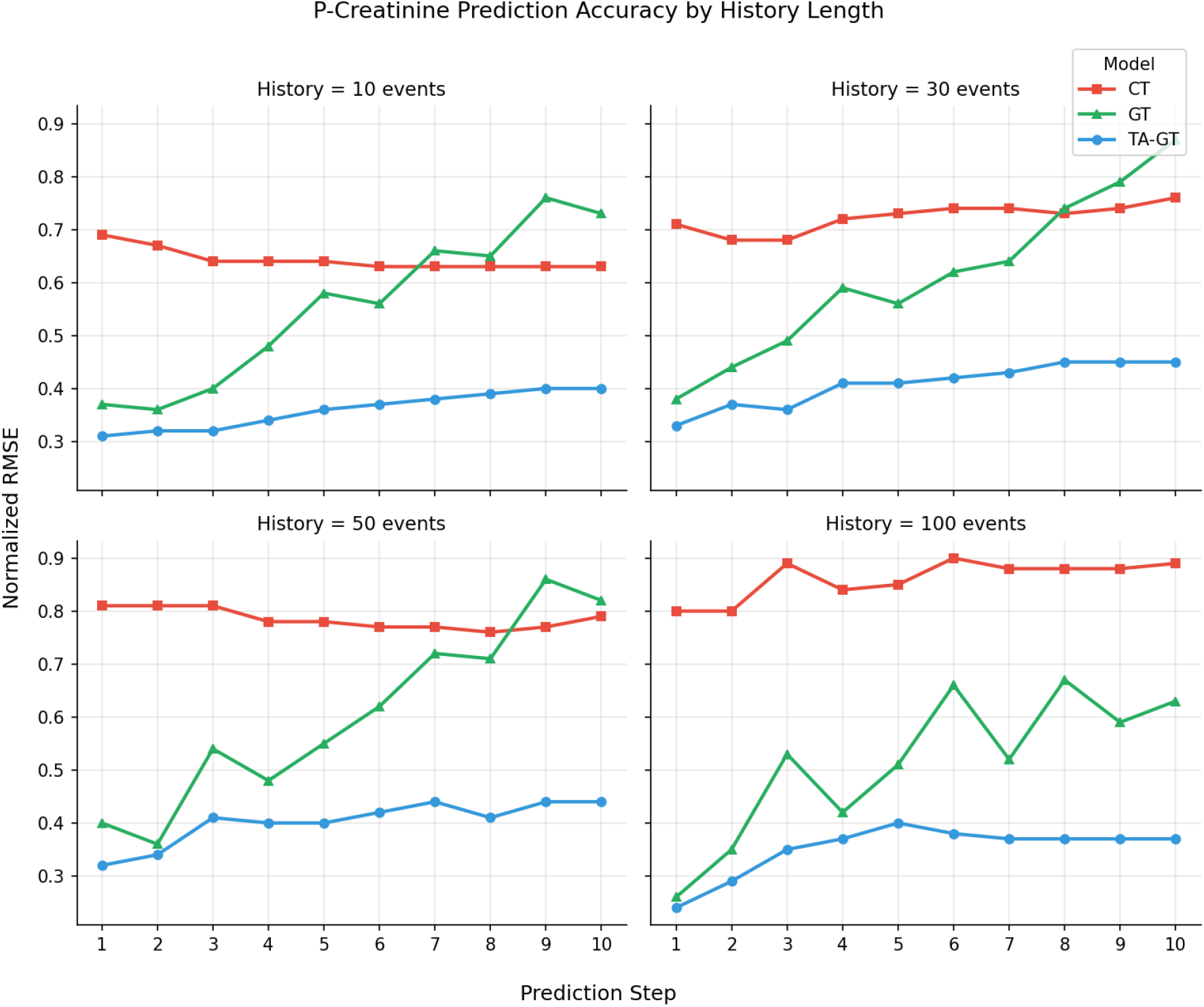
Plasma creatinine prediction accuracy (normalized RMSE) vs. prediction step for different history lengths. TA-GT (blue) maintains stable error across horizons while GT (green) and CT (red) degrade. Lower is better. Prediction starts after the given timesteps of history.

Clinical decision support systems require not only accurate point predictions but also reliable uncertainty estimates. Appendix C compares prediction intervals from MSE and heteroscedastic NLL training by assessing using two criteria: calibration, which measures the agreement between nominal confidence and empirical coverage, and sharpness, which compares the narrowness of interval widths.

#### 5.2.2 Effects OF IRREGULAR SAMPLING

Prediction accuracy depends on the elapsed time between subsequent samples. Figure 4 shows TA-GT’s one-step prediction error as a function of elapsed time since the last measurement of different laboratory markers. Predictability varied by covariate: Potassium showed consistently the largest error regardless of elapsed time. We observed that mean corpuscular hemoglobin (MCH) was highly predictable at short horizons, with error increasing as the temporal gap widened. In contrast, plasma creatinine exhibited greater predictability over longer intervals. A plausible explanation for these patterns is that longer measurement gaps in routine care often correspond to clinically stable outpatients, for whom creatinine dynamics are comparatively slow, whereas shorter gaps typically occur in inpatient settings with greater physiological volatility.

**Figure 4:**
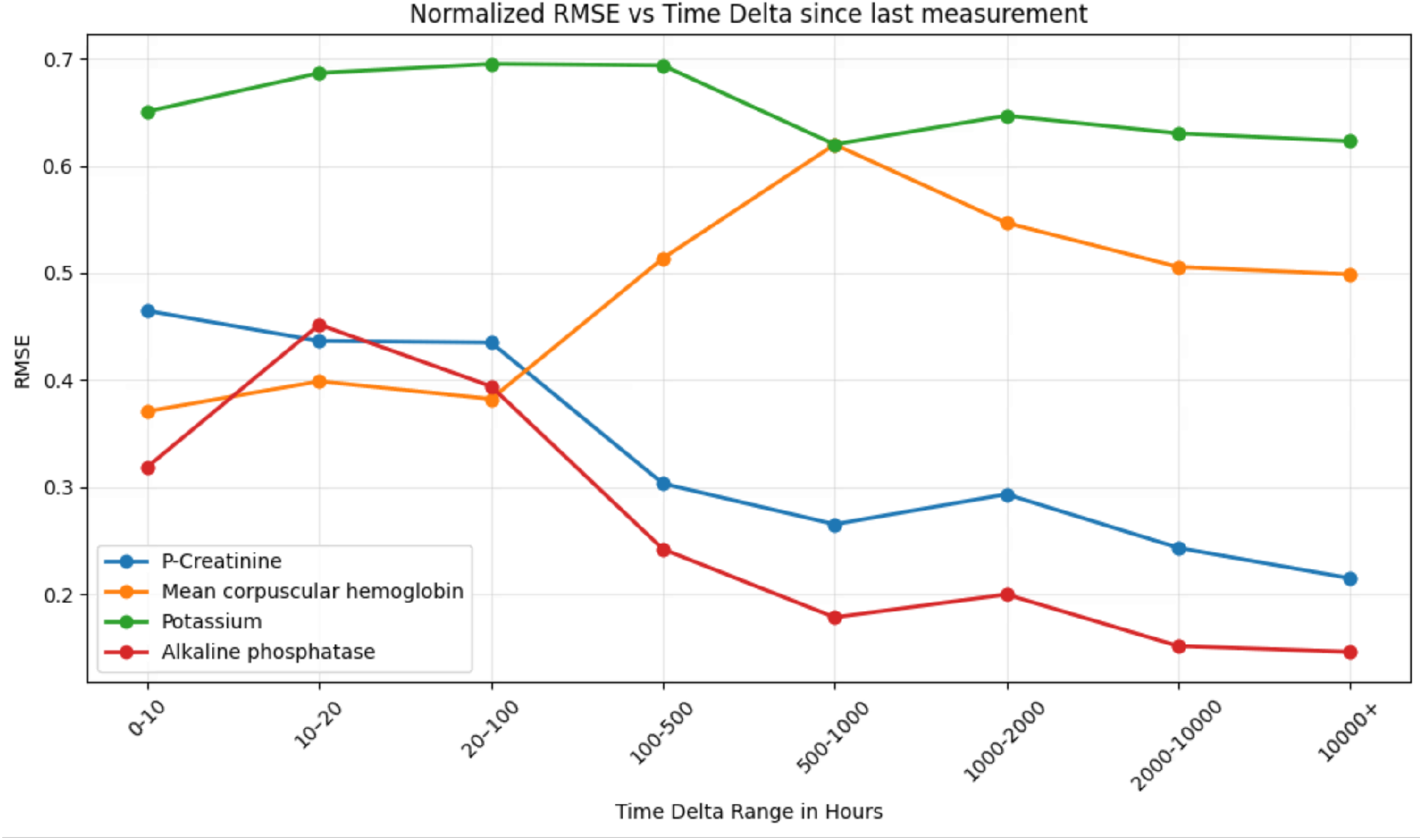
TA-GT’s one-step-ahead normalized RMSE vs. elapsed time since last measurement for four laboratory markers. These patterns reveal how each biomarker’s predictability depends on measurement frequency: stable markers like plasma creatinine remain predictable over long gaps, whereas volatile markers like Potassium are inherently harder to predict regardless of timing.

#### 5.2.3 Population-Level Trajectory Prediction

We evaluated population-level trajectory prediction by predicting plasma creatinine values following the first administration of propionic acid derivatives (M01AE). Starting from the moment of treatment administration, we predicted up to the last recorded measurement for each patient. To summarize results over time, predictions and ground-truth observations were aggregated into bins on a logarithmic time scale, where each bin contains measurements that fall after the previous bin boundary and up to the current one. This binning strategy provides denser coverage of the immediate post-treatment period while still capturing longterm dynamics.

As shown in Figure 5, TA-GT closely follows the true population-level plasma creatinine trajectory throughout the entire prediction horizon, including at later time points where observations become sparse. In contrast, both GT and CT tended to overpredict creatinine levels, with their estimates diverging from the observed trend particularly at longer time scales. This demonstrates TA-GT’s ability to capture the direction and magnitude of population-level treatment effects over an extended and irregular prediction window.

**Figure 5:**
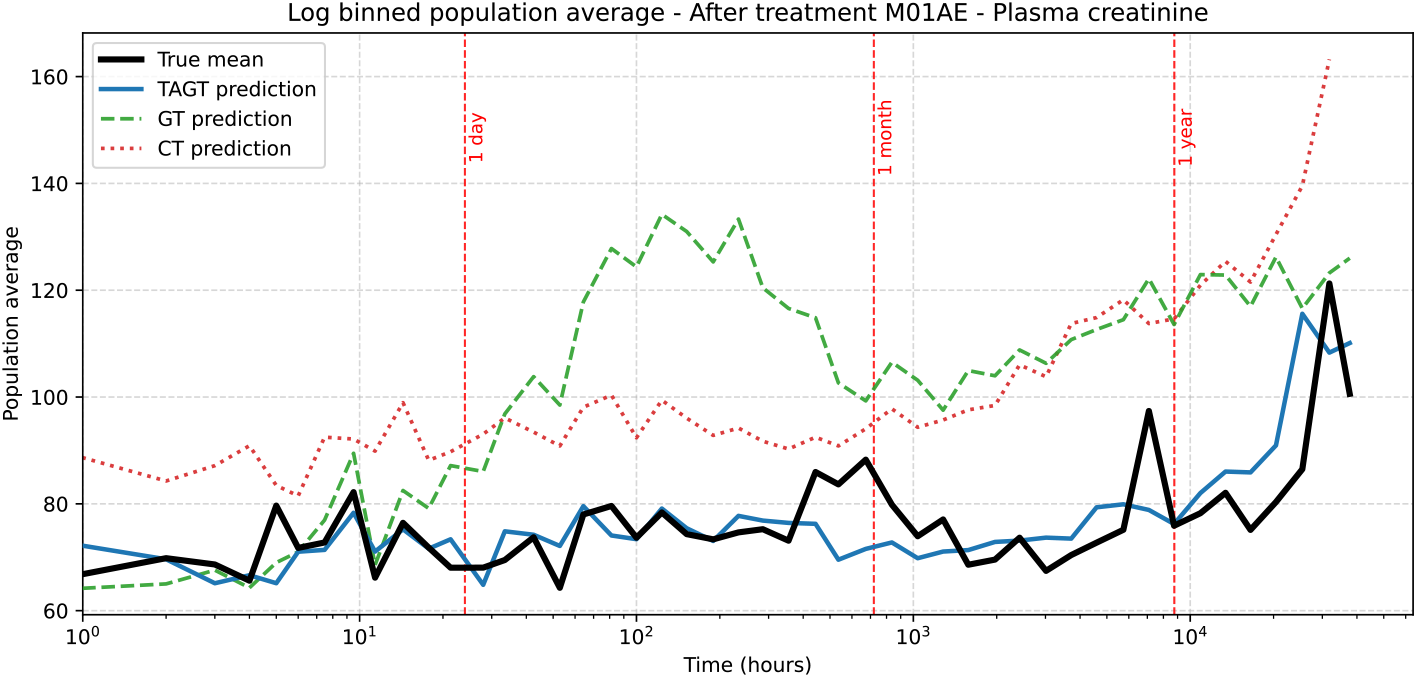
Population-level mean plasma creatinine trajectory following M01AE treatment, aggregated into logarithmic time bins after treatment administration. Each bin averages values observed after the previous bin boundary and up to the current one. Time 0 is aligned to first treatment administration. TA-GT (orange) follows the observed trend (blue) across the full prediction horizon, while GT and CT overpredict creatinine levels.

### 5.3 Ablation Study

To validate our architectural contributions, we conducted an ablation study on the realworld dataset. We systematically removed key components from the full TA-GT model and measured the impact on ten-step-ahead prediction RMSE for plasma creatinine, using a total history of 50 events prior to the medication. The evaluated modifications were: (i) removing the measurement mask embeddings 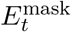 (Eq. 4), which encode which covariates were observed at each time step; (ii) removing the next time-delta conditioning 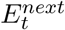 (Eq. 11), which informs the model about the time until the next observation; and (iii) removing the Time Relation Estimation bias *R*^∗^ (Eq. 5) from the attention scores. As shown in Table 3, each component played a significant role, with the removal of the **measurement mask** causing the greatest error increase from 0.40 to 0.51.

**Table 3:** Performance degradations due to ablations with ten-step-ahead prediction of plasma creatinine level based on history of 50 events.

| Model Modification | RMSE (normalized) | % Change |
| --- | --- | --- |
| **Full TA-GT** | **0.40** | — |
| w/o Time Relation (TRE) | 0.45 | +12.5% |
| w/o Next Time-Delta ($\Delta$) | 0.50 | +25.0% |
| w/o Measurement Mask | 0.51 | +27.5% |

## 6 Discussion

Our results demonstrate that integrating attention-level temporal awareness with G-computation improves long-term counterfactual prediction for irregular clinical time series. The proposed model consistently surpassed adapted state-of-the-art baselines in both synthetic evaluations and a large, real-world EHR dataset. TA-GT exhibited stable performance across a wide range of prediction horizons, whereas the baseline models showed rapid performance degradation. Notably, TA-GT was the only model that accurately captured population-level treatment effects following M01AE administration, both at short-term and long-term prediction horizons

The real-world evaluation also revealed substantial heterogeneity in predictability across laboratory markers. As shown in Figure 4, plasma creatinine remained predictable even over long measurement gaps, whereas potassium exhibited high prediction error at all horizons. Mean corpuscular hemoglobin fell between these extremes, showing strong short-term predictability that diminished over longer intervals. These findings indicate that prediction reliability depends fundamentally on the specific biomarker rather than being uniform across laboratory tests. In practice, stable markers may support longer-horizon treatment planning, whereas more volatile markers are most informative at short horizons or when combined with closer monitoring.

Although our real-world evaluation relies on factual prediction accuracy, since true counterfactual outcomes are inherently unavailable in observational data, the G-computation framework provides principled causal adjustment. Our formulation addresses the challenge of informative measurement timing in EHRs by reframing the causal estimand as a joint intervention on both treatments and future measurement timings under a *Measurement Timing Ignorability* assumption. While the model conditions on a high-dimensional history of covariates and measurement masks to approximate this, explicitly modeling the observation process alongside the clinical state remains a direction for future work. As with all observational causal inference approaches, the standard assumptions of no unmeasured confounding and positivity apply; consequently, the model is best suited for evaluating realistic treatment and monitoring strategies that are well-represented in the training data, rather than arbitrary out-of-distribution interventions.

In conclusion, our results highlight the importance of explicitly encoding temporal irregularity within the attention mechanism for clinical time series, where the observation process itself carries diagnostic information. The combination of the time relation estimation bias, measurement mask embeddings, and next-time-delta conditioning allows TA-GT to jointly reason about *what* was measured, *when* it was measured, and *how long* until the next observation. Furthermore, TA-GT equipped with faithful heteroscedastic NLL training produces well-calibrated and sharp uncertainty estimates without sacrificing predictive performance, enabling clinicians to distinguish high-certainty predictions from those where the model appropriately signals uncertainty. These properties make TA-GT a practical tool for retrospective treatment evaluation and clinical hypothesis generation, supporting the design of treatment strategies in settings where randomized controlled trials are infeasible or unethical.

## Data Availability

Sensitive patient data from the HUS Helsinki University Hospital cohort cannot be distributed according to Finnish national regulations and EU legislation. All analyses were conducted within the secure HUS Acamedic platform. Synthetic data generation code and model implementation will be made publicly available upon publication.

## Data Availability

The MD-JoPiGo framework and its validation testing datasets are openly available at https://github.com/ZheqingZhu/mdjopigo. The empirical datasets used in this study (NCCTG lung cancer and Moertel colorectal cancer) are publicly accessible via the R 'survival' package (https://CRAN.R-project.org/package=survival).

## 7 Author Contributions

G.H.: Methodology, Implementation, Experiments, Writing, A.H.: Data curation, Writing, Reviewing, K.S.: Writing, Reviewing, S.S.: Supervision, Writing, Reviewing, R.R.: Supervision, Writing, Reviewing, Funding acquisition, M.K.: Conceptualization, Supervision, Writing, Reviewing, Funding acquisition

## 8 Acknowledgements

We acknowledge funding from the Helsinki University Hospital VTR funds (grant no. TYH2025370 to R.R) and from Business Finland (grant 4911/31/2024 to R.R.). The authors declare no financial or non-financial competing interests.

## 9 Ethical considerations

Following national and EU legislation, no ethical permission was required for retrospect registry data and thus the study was based on approval of HUS Helsinki University Hospital (permission HUS/355/2025, HUS/223/2023). The data storage and analysis were conducted within a secure analytics platform (HUS Acamedic), ensuring patient confidentiality and compliance with Finnish Medical Research Act for the secondary use of medical records (552/2019).

## 10 Data Availability

Sensitive patient data from the HUS cohort cannot be distributed according to national regulations. Synthetic data generation codes are available at [upon publishing].

## 11 Code Availability

The implementation of the proposed model is available at the public repository [upon publishing] under MIT licence.

## Appendix A. Baseline Model Modifications

The baseline models, GT and CT, were originally proposed with the assumption of regularly sampled data. To ensure a more fair comparison on our irregular datasets, we implemented several adaptations. These modifications were designed to remain as faithful as possible to the original papers while enabling them to handle irregular time intervals and informative missingness. The adaptations differed between the synthetic and real-world datasets due to their distinct temporal characteristics.

### A.1 Synthetic Dataset Adaptations

#### Time Encodings

For GT, we utilized absolute sinusoidal time encodings. However, instead of using token order, we provided the integer day counts as the input to the time embedding function. For CT, we retained its original relative positional encoding mechanism but incorporated time information by adding the day indices into the encoding procedure.

#### Next Time-Delta Conditioning

This feature was not present in the original models but is used by our proposed TA-GT. To ensure a fair comparison, we added this information to both baselines. The next time-delta was embedded using a small MLP and added to the hidden state before the final projection heads.

### A.2 Real-World Dataset Adaptations

#### Time Encodings

For GT, the large and sparse time gaps in the HUS data made absolute sinusoidal encodings unstable. We replaced them with Time2Vec, which serves as a generalization of sinusoidal encodings but with learnable parameters, offering more flexibility. For CT, we again kept its relative positional encoding. To handle the wide-ranging time differences (from hours to years), we applied the time differences on a logarithmic binned scale to ensure that both small and large gaps remained distinguishable.

#### Next Time-Delta Conditioning

This was implemented identically to the synthetic data adaptation.

#### Measurement Mask

Neither original baseline explicitly models the missingness of covariates. For fairness, and mirroring the architecture of TA-GT, we applied a measurement mask embedding to the hidden state of both models. This allows them to encode information about the presence or absence of measurements at a given timestep.

## Appendix B. Learning Gaussian Distributions with Faithful NLL

To properly quantify aleatoric uncertainty, we aimed to predict a full probability distribution for each covariate rather than just a single point estimate. The standard MC simulation, which samples from empirical residuals, assumes the data’s noise is **homoscedastic**, which means that the variance of the error is constant and does not depend on the patient’s state or the time gap between measurements.

Shifting from a Mean Squared Error (MSE) loss to a Negative Log-Likelihood (NLL) loss allows the model to learn this **heteroscedastic** variance by outputting the parameters of a Gaussian distribution 𝒩 (*µ*(*H*), *σ*^2^(*H*)) based on the patient’s history *H*. However, optimizing with a *vanilla* NLL loss can degrade the accuracy of the mean prediction (*µ*). The model may learn to “explain away” difficult-to-predict samples by simply increasing their variance (*σ*^2^), which satisfies the NLL loss but harms the predictive accuracy of the mean.

To solve this, we used **Faithful Heteroscedastic Regression** [15]. This method modifies the loss function by adding an MSE term to the NLL loss. Also, it uses **stopped gradients** to decouple the optimization of the mean and the variance. The gradients from the NLL loss are stopped from influencing the mean-prediction heads, while gradients from the MSE loss are stopped from influencing the variance-prediction heads. This allows the model to learn an accurate mean prediction while separately learning a well-calibrated variance, ensuring that the two objectives do not interfere.

Table B1 shows the 1-to-10-step-ahead RMSE for plasma creatinine (using a 100-event history). It is nearly identical for the TA-GT model trained with Faithful NLL and the standard TA-GT model trained with MSE.

**Table B1:** Comparison of plasma creatinine 10-Step-Ahead Normalized RMSE (History=100) by Loss Function.

| Model | Step 1 | Step 2 | Step 3 | Step 4 | Step 5 | Step 6 | Step 7 | Step 8 | Step 9 | Step 10 |
| --- | --- | --- | --- | --- | --- | --- | --- | --- | --- | --- |
| TA-GT (MSE) | 0.26 | 0.30 | 0.35 | 0.37 | 0.39 | 0.39 | 0.40 | 0.41 | 0.42 | 0.43 |
| TA-GT (NLL) | 0.26 | 0.30 | 0.35 | 0.37 | 0.39 | 0.39 | 0.40 | 0.40 | 0.42 | 0.43 |

## Appendix C. Calibration of Predictive Uncertainty

### C.1 Calibration and Uncertainty Quantification

For clinical decision support, a model’s predictions must be reliable, and its uncertainty estimates must be well-calibrated. We evaluate the quality of prediction intervals from G-Transformer with residual sampling, TA-GT with residual sampling, and TA-GT with faithful NLL sampling from the predicted distributions. We assess calibration using coverage and the Winkler score [16] to evaluate both the reliability and informativeness of the intervals.

#### Coverage

This measures the frequency that the true, observed outcome falls within the model’s predicted confidence interval (e.g., a 90% interval should ideally contain the true value 90% of the time).

#### Winkler Score

Coverage alone can be misleading, as a model can achieve high coverage by producing overly wide and uninformative intervals. The Winkler score penalizes both wide prediction intervals and predictions falling outside of the interval. The Winkler score is defined as:

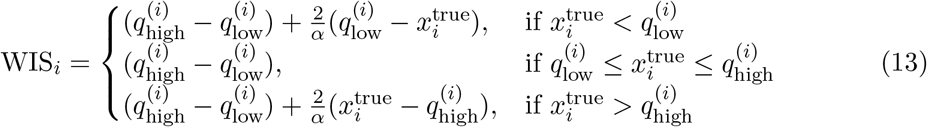

Figure C1 shows the overall calibration plot, where the dashed line represents ideal calibration. All models are reasonably well-calibrated. As shown in Figure C2, TA-GT (NLL) achieves the lowest (best) Winkler score across all 1-10 prediction steps by slightly beating residual sampling. While the standard GT model produces intervals with high coverage by creating wider intervals, leading to a higher Winkler score.

**Figure C1:**
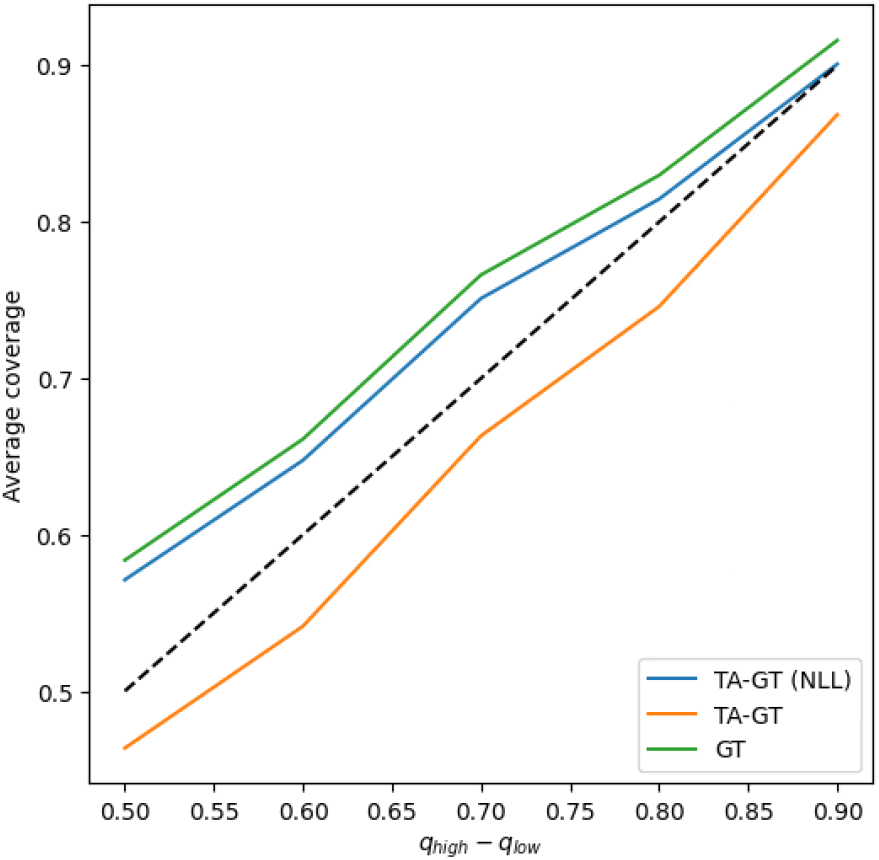
Overall calibration plot. The dashed line is ideal. All models are reasonably calibrated, with TA-GT (NLL) staying close to the ideal.

**Figure C2:**
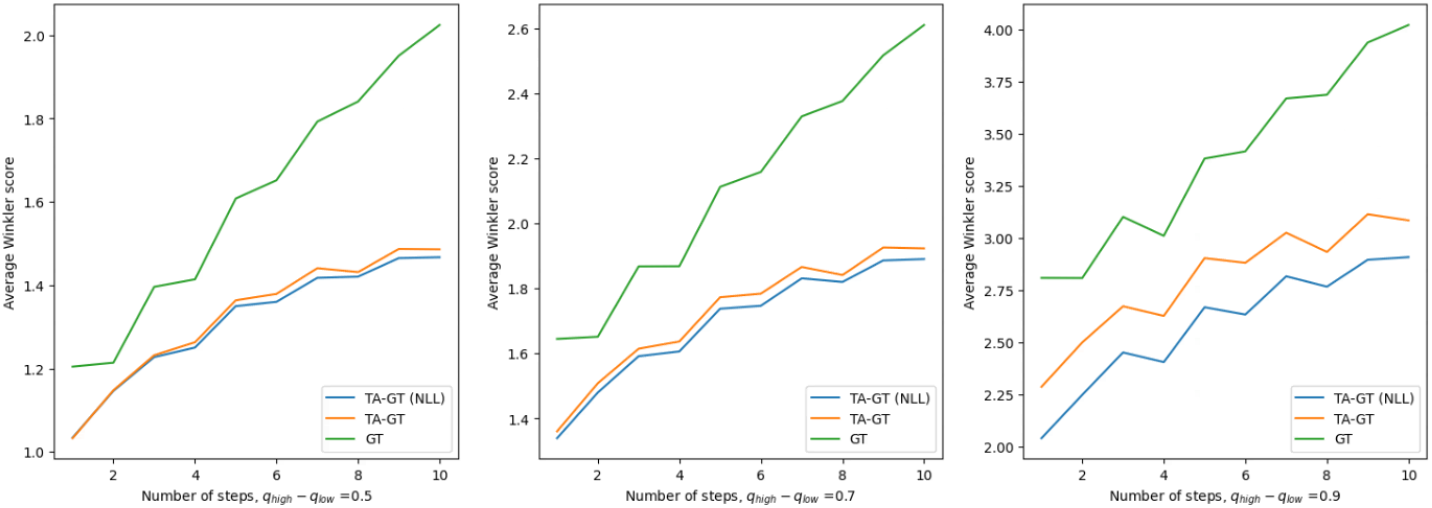
Winkler interval scores for 1-10 step-ahead predictions at 90% confidence. **TA-GT (NLL)** achieves the lowest (best) score, indicating the most informative and well-calibrated uncertainty estimates.

## Appendix D. HUS dataset

Table D1 lists all types of continuous laboratory measurements included in the dataset used in our experiments. The variables are presented with their Finnish names alongside their English equivalents.

**Table D1:**
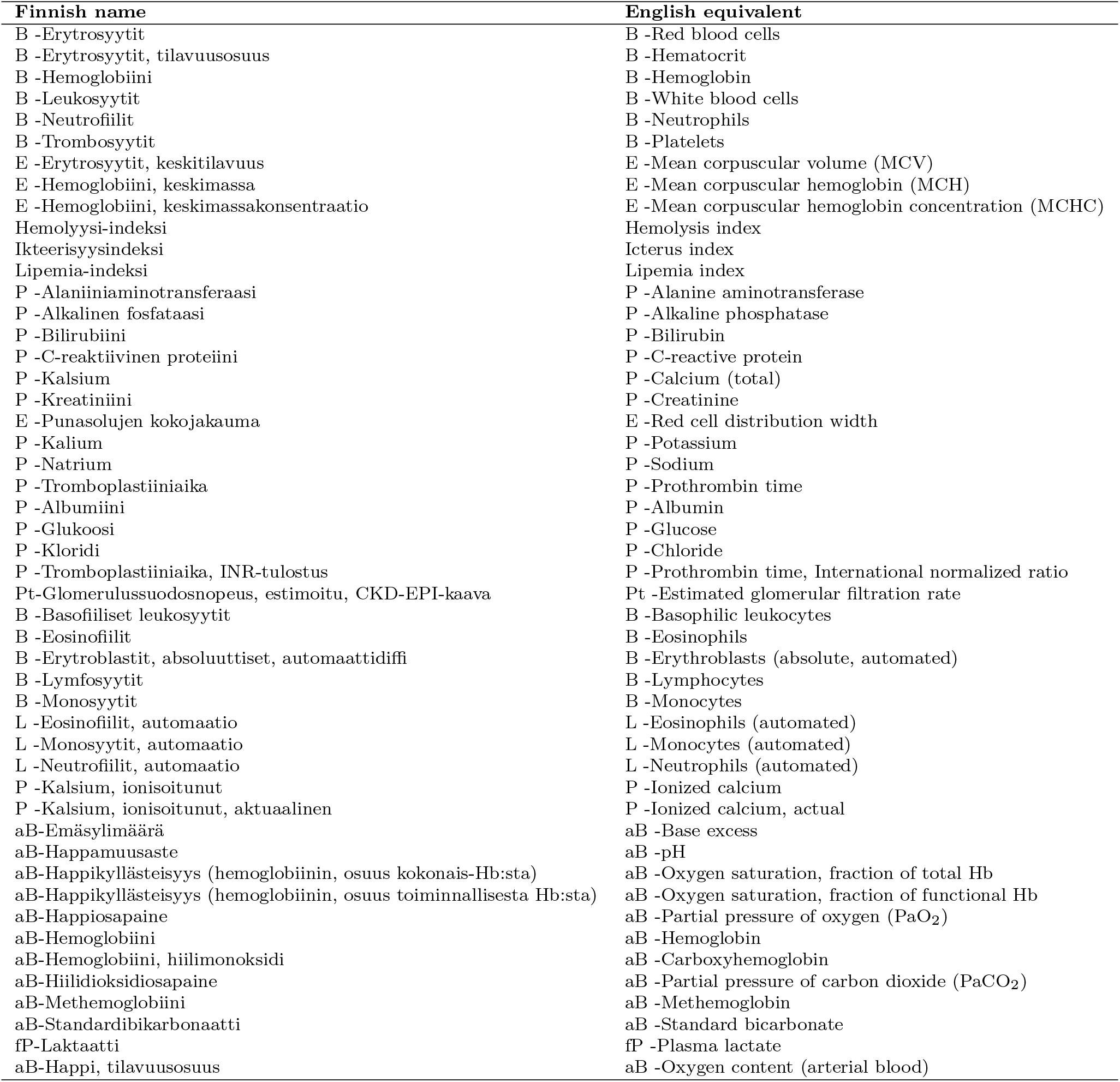
Laboratory variables included in the real-world dataset.

Specimen prefixes used in Table D1: P (plasma), B (blood), E (erytrocyte), U (urine), L (leukocyte differential count), fP (fasting plasma), aB (arterial blood), S (serum). Hemolysis index, Icterus index, and Lipemia index are sample-quality indices.

**Figure D1:**
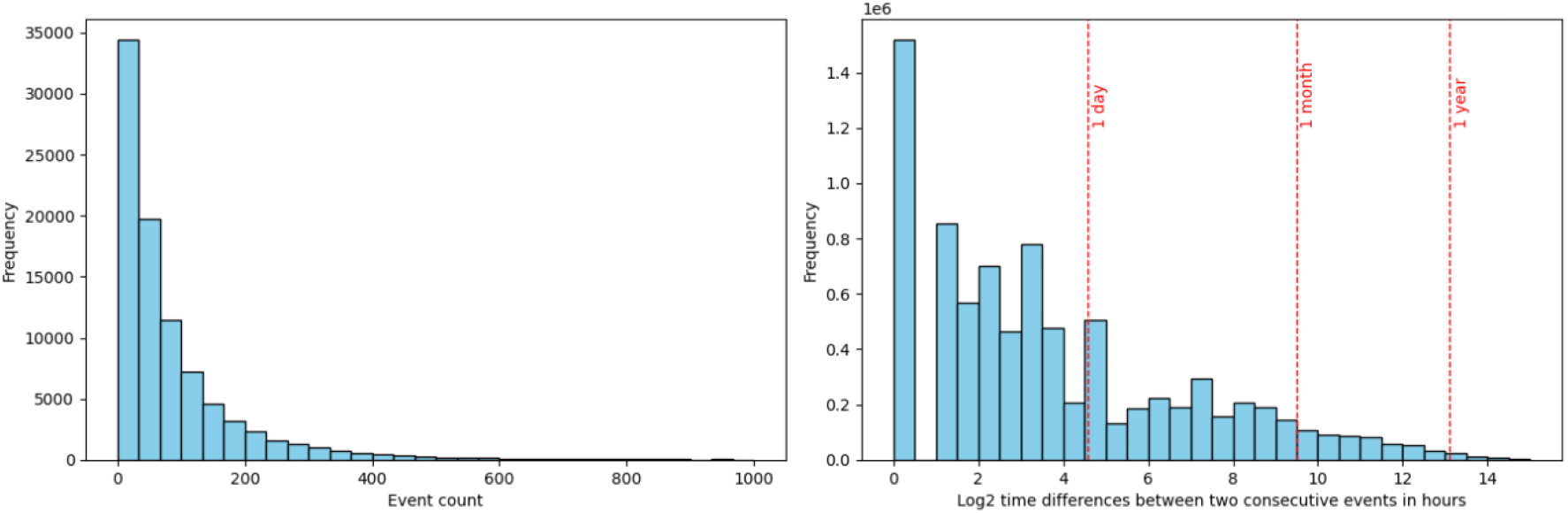
The distribution of medical events per patient in bins by their event count, and time differences between two consecutive timestamps

**Figure D2:**
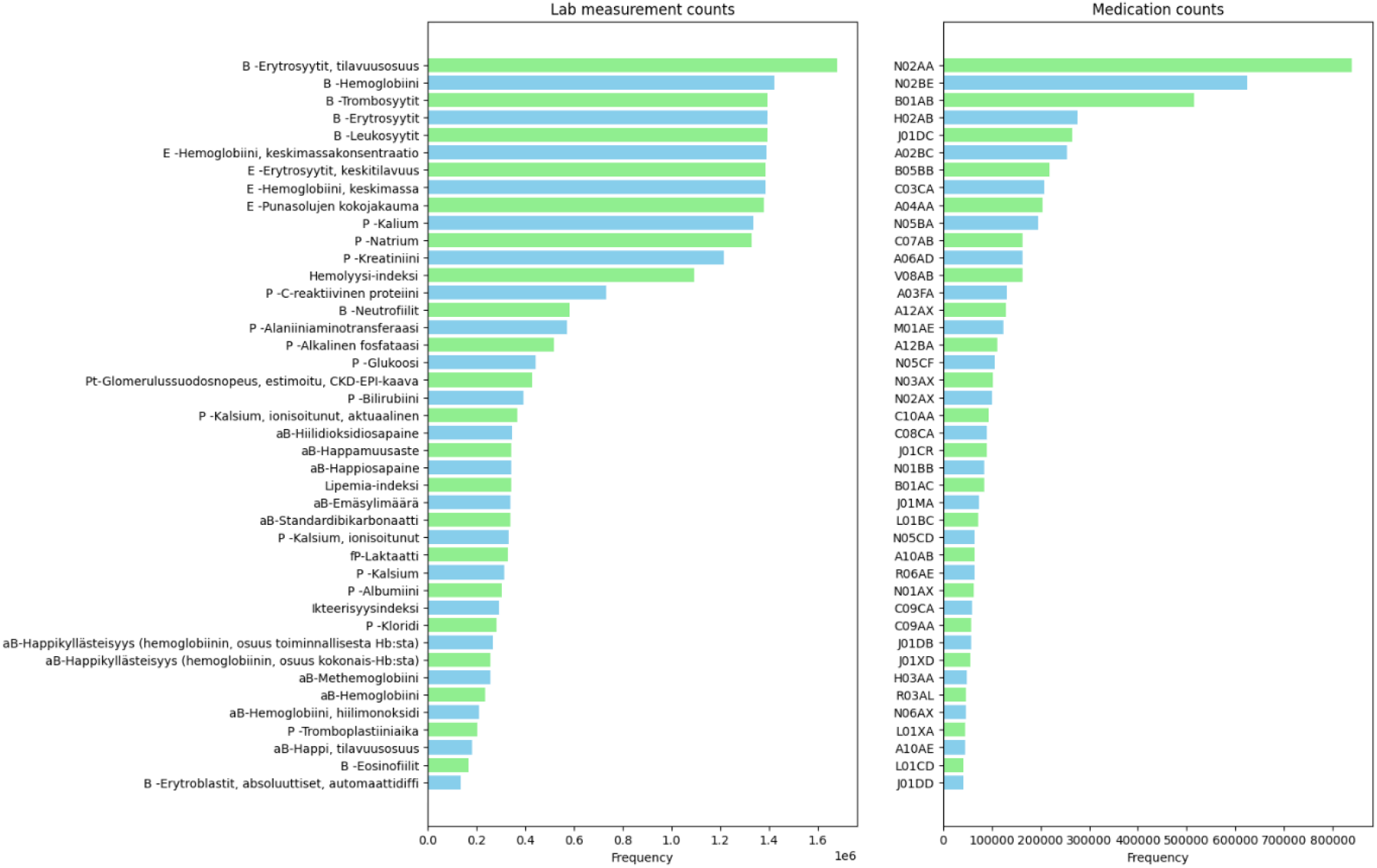
The distribution of all laboratory measurements and medications in real-world data.

## Appendix E. Implementation Details

All models were implemented in PyTorch and trained on an NVIDIA T4 GPU with 16GB of VRAM. For faster attention calculation we utilized FlashAttention for a more memory-efficient calculation of scaled dot-product attention.

Models were trained using AdamW optimizer with a weight decay of 0.01, and we employed a learning rate scheduler with a step size of 5 and a gamma of 0.1. Training ran for a maximum of 150 epochs, with early stopping based on the validation loss (patience of 5 epochs) to prevent overfitting. Dropout with rate of 0.1 was used across all network layers.

For the Causal Transformer baseline, we followed the original training procedure, including an adversarial confusion loss (*α* = 0.01) with an exponential rise and an exponential moving average of parameters (*β* = 0.99).

Optimal model hyperparameters were determined using a random grid search, with the final configuration for each model selected based on the lowest RMSE on the validation set.

